# The Effects of Reducing pH with a Bioresorbable Synthetic Matrix on the Diabetic Foot Ulcer Microenvironment

**DOI:** 10.64898/2026.09.08.26362071

**Authors:** Lauren Lie, Simon Tabchi, Lindsay Kalan

## Abstract

**Objective:** Non-healing diabetic foot ulcers (DFUs) stall in an alkaline and pro-inflammatory microenvironment that delays healing and increases the risk of complications. Since an acidic pH has been correlated with healing, wound acidification has become an appealing strategy to perturb the DFU microenvironment and promote wound closure. While reducing wound pH has shown promising results, the effects of pH on inflammation and the microbiome have not been well characterized. Along with inflammation, microbial communities are critical to the wound microenvironment and are associated with healing trajectories. We aimed to explore the longitudinal effects of altering pH on the DFU microbiome and inflammatory profile.

**Approach:** 10 subjects, each with a chronic DFU, were treated with a polyglycolic acid/ poly(lactide-co-caprolactone) bioresorbable matrix over a 4-week period, and changes in wound pH, volume, inflammation, and microbiome were evaluated every 14 days.

**Results:** Wounds decreased significantly in pH (p<0.001) and volume (p<0.05). Wound-specific changes to both bacterial and fungal communities occurred, with alkaline wounds increasing in bacterial diversity as pH decreased. Wound size and baseline pH were found to be associated with microbial composition, and larger wounds were elevated in pro-inflammatory chemokines.

**Innovation:** To capture the complete bacterial and fungal presence, 16S and ITS amplicon sequencing was used to assess community-level changes to wound acidification.

**Conclusions:** The relationship between greater wound size, microbial composition, alkalinity, and inflammation demonstrates their interconnected role in the DFU microenvironment and wound severity. This work supports that wound acidification may have the potential to perturb microbial communities with a greater effect on alkaline DFUs.

## Introduction

One of the most serious complications associated with diabetes is the development of a diabetic foot ulcer (DFU). DFUs frequently progress to more serious complications such as infection and lower-limb amputation, resulting in poor prognoses and diminished quality of life.^1^ In addition to patient impact, DFUs have a considerable economic burden on healthcare systems, which is projected to grow as global diabetes prevalence increases.^2^ In DFUs, healing is hindered by diabetes-related comorbidities, such as neuropathic and vascular disease, and an ineffective immune response that stalls the wound in a chronic low-grade pro-inflammatory state.^1,3^ Wound deterioration is further exacerbated by imbalances within the complex microenvironment, encompassing oxygenation, inflammation, pH, and the microbiome.^3–7^ Current standard of care emphasizes glycemic control, removal of necrotic tissue, infection prevention, and offloading.^8^ However, despite these interventions, only 30-40% of DFUs heal after 3 months of treatment and almost a quarter remain unhealed at 12 months^1,9^ Consequently, emerging therapeutic strategies have shifted towards modulating the DFU microenvironment to promote conditions conducive to healing.^4^

Soon after ulceration, bacteria and fungi can colonize the wound and are hypothesized to form a polymicrobial community referred to as the DFU microbiome. This includes skin commensals, and while normal skin flora might stimulate the immune system for antimicrobial defense and tissue regeneration, dysbiosis with pathogenic species of microorganisms can impair wound healing.^7,10,11^ In non-healing DFUs, studies have shown an association with reduced bacterial diversity compared to healthy skin and increased proportion of anaerobic and pathogenic bacteria such as *Enterobacter* and *Staphylococcus aureus*.^12,13^ While much less is understood about fungal presence in chronic wounds, greater fungal diversity has been associated with increased necrotic tissue and wound deterioration.^14^ Temporal community stability has also been associated with poor outcomes and is hypothesized to reflect inefficient clearance of bacteria by the immune system. Conversely, a more dynamic and changing microbiome has been associated with healing outcomes.^15^ Thus, treatment that can perturb the chronic wound microbiome and restore a balanced community found in healthy skin may help improve DFU outcomes.

Wound pH has long been hypothesized to be implicated in wound healing and has increasingly been incorporated into treatment strategies. In acute wounds, pH begins slightly alkaline, near 7.4, and progressively decreases to a pH much closer to acidic healthy skin over the healing trajectory. However, DFUs can stall in an alkaline/basic microenvironment, reaching a pH as high as 9, contributing to hypoxia and inflammation, which ultimately exacerbates tissue damage.^6^ Thus, an acidic pH has been correlated with healing, whereas chronic non-healing wounds are associated with a basic microenvironment.^16,17^ This has resulted in recent studies exploring the effects of acidification through acid-releasing materials. In murine wound models, reducing pH has shown promising results in increasing oxygenation and re-epithelialization rates.^18–20^ Acidification has also been suggested to reduce inflammation by shifting macrophage polarization towards an M2 anti-inflammatory phenotype, potentially minimizing excessive inflammatory signalling.^21,22^ Together, these effects may impact bacterial and fungal communities by altering their growth.^23^ Indeed, some pH modulating dressings have been shown to slow the growth and proliferation of *S. aureus* and *Escherichia coli in vitro*.^24,25^ In fungi, acidic environments can also induce a yeast phenotype by inhibiting hyphal growth, a virulence trait that permits tissue invasion.^26,27^ However, it is still not well understood how reducing wound pH affects overall microbial community structure over time.

Here, we conducted a pilot study to test the effect of a polyglycolic acid/ poly(lactide-co-caprolactone) (PGA/PLCL) bioresorbable matrix on the DFU microbiome. The synthetic matrix is created via electrospinning PGA and PLCL polymers that slowly release glycolic, lactic, and caproic acids into the wound bed as it degrades over a 2-week period. These properties are designed to create a localized acidic microenvironment and promote tissue regeneration. To assess the changes in wound pH, immune profile, and microbiome, 10 subjects with chronic DFUs were recruited and received application of the matrix over a 4-week study period, including weekly secondary dressing changes and matrix reapplication every 14 days. Changes in wound pH were assessed throughout treatment and compared to baseline, and amplicon sequencing was performed to analyze shifts in bacterial and fungal communities. Trends associated with the DFU immune profile and clinical wound features were also evaluated.

### Innovation

While the growth and functional activity of microorganisms are altered by environmental pH, it is not well understood how the wound microbiome is affected by altering wound pH.^23^ Culture based and targeted PCR methods have been used to explore the association between wound pH and the presence of bacterial taxa; however, these methods often underestimate microbial diversity and have excluded fungi.^28–30^ Here, we treated 10 chronic DFUs with a PGA/PLCL matrix and used 16S and ITS amplicon sequencing to characterize the bacterial and fungal presence and evaluate changes in the complete wound microbiome over time as local pH shifts.

### Clinical Problem Addressed

Currently, standard of care results in low healing rates with only 30-40% of DFUs healing after 3 months of treatment. Prolonged ulceration substantially increases the risk of complications including infection and amputation, perpetuating high treatment costs.^9^ Non-healing DFUs exist within a stalled alkaline and pro-inflammatory microenvironment that can lead to increased bacterial growth and delay progression through the phases of wound healing. Since an acidic pH has been correlated with wound healing, recent therapeutic strategies have used wound acidification to disrupt the microenvironment and generate optimal healing conditions.^6^ While some studies have explored the effects of reducing wound pH on healing and inflammation, few have assessed how altering pH affects the complex community of microorganisms within the wound bed. Microbial community structure is a core component to the wound microenvironment and are associated with wound outcomes.^12^ To better understand the effects of altering wound pH, we assessed if the DFU microbiome and immune profile are altered by treatment with a PGA/PLCL synthetic matrix.

## Materials and Methods

### Study design and sample collection

For this pilot study, 10 adults, each with a chronic DFU, were recruited for treatment with a PGA/PLCL matrix. Eligible DFUs were non-infected, partial- or full-thickness ulcers that failed to achieve a minimum of 50% area reduction after 4 weeks of standard of care, comprising surgical debridement, dressings to maintain a moist wound environment and manage exudate, off-loading, vascular assessment, and infection and glycemic control. Subjects visited the PA Foot & Ankle Associates clinic (Lehigh Valley, PA, USA) weekly for 4 weeks (5 visits). Patient history and baseline data were collected at visit 1, and progress through treatment was assessed at visits 2 through 5. At visits 1, 3 and 5, sample collection and sharp debridement was performed before application of the PGA/PLCL matrix, except in 2 cases where wounds had healed, precluding the need for debridement at visit 5. Wound pH was measured using non-bleeding pH indicator strips. Sterile foam tip and nylon flocked swabs moistened with sterile NaCl (0.15M) solution were used to sample deep wound fluid from the wounds centre. For microbiome analysis, swabs were also collected from intact skin 5-10 cm from the wound and from clinic air for a negative control. All swabs were preserved in a DNA/RNA shield (Zymo Research, Irvine, CA, USA) at -40°C until further processing. Between sample collection weeks at visits 2 and 4, wounds received standard of care without debridement.

Swabs collected for immune marker analysis were processed by RayBiotech Inc. (Peachtree Corners, GA, USA). Enzyme-Linked Immunosorbent Assay (ELISA) was used to quantify the concentrations of 40 immune markers. Only analytes with a minimum of 80% of values within the lower and upper limits of quantitation were further analysed and concentrations were normalized to the total protein concentration within each sample.

Swabs for DNA extraction (wound n=28, intact skin = 28, negative control = 18) were processed alongside an oral microbiome mock community as a positive control since skin and oral sites are known to share similar taxa. To process all wound fluid, swabs were centrifuged in a filter tube insert for 1 minute at 21 000 x g. Samples were then processed on a vortex genie 2 (Scientific Industries, Bohemia, NY, USA) for 40 minutes of continuous bead beating in bead bashing tubes (Zymo Research, Irvine, CA, USA) and centrifuged at 10 000 x g for 1 minute. Total DNA was then isolated by following the ZymoBIOMICS DNA miniprep kit (Zymo Research, Irvine, CA, USA) as per the manufacturer’s instructions. DNA was eluted in 50uL of elution buffer and quantified using a Qubit 2.0 Fluorometer (Life Technologies, Carlsbad, CA, USA) with the Qubit dsDNA broad range assay kit (Life Technologies, Carlsbad, CA, USA). All data were recorded in an electronic laboratory notebook with the Lab Archives platform.

### 16S rRNA and ITS amplicon sequencing

Paired end amplicon sequencing of the V3-V4 region of the 16S rRNA bacterial gene and internal transcribed spacer 1 (ITS1) fungal region was completed on an Illumina MiSeq platform by the Genomics Facility at McMaster University. The resulting bacterial and fungal reads were processed separately but followed a similar quality control process. 16S V3-V4 raw reads were first processed by fastp (v0.23.2) to remove low quality reads and trim adapters and poly-g tails.^31^ Changes in sequence quality were assessed and visualized with FastQC (v0.11.9) and MultiQC (v1.12).^32,33^ Further quality filtering was performed within a Quantitative Insights Into Microbial Ecology 2 (QIIME2) environment (v2025.10).^34^ Primer sequences were removed with Cutadapt (v5.1).^35^ DADA2 (v1.30.0) trimmed low quality ends to 260 and 230 nucleotides for the forward and reverse reads, respectively, as well as denoised, merged paired end reads and removed chimeras.^36^ Vsearch (v2.22.1) clustered the processed reads into operational taxonomic units (OTUs) based on 97% sequence similarity.^37^ Taxonomic assignment was performed by a Naïve Bayes classifier pre-trained on the Genome Taxonomy Database r220 full length sequences (v220.0).^38^ Since ITS1 regions have variable lengths, reads processed by fastp were trimmed to known ITS1 start and stop sequences with ITSxpress (v2.1.4).^39^ DADA2 was used to denoise, merge paired end reads and remove chimeras. Reads shorter than 100 base pairs were removed and the remaining quality filtered reads were clustered into 97% OTUs. BLAST+ local alignment between ITS1 OTUs and reference reads from UNITE+ INSD database for fungi (v19.02.2025) was used for classification.^40^

### Statistical analysis of the microbiome and clinical variables

All statistical analyses were conducted using the R programming language and the Tidyverse (v0.99.6) and dplyr packages (v1.1.4) were used to prepare and organize data for analysis.^41–43^ The microbiome data were analysed using the phyloseq package (v1.52.0).^44^ Top abundant OTUs not classified to the genus level were queried against the NCBI BLAST+ 16S rRNA and ITS databases and re-classified to the taxonomic level where top alignments reached agreement. Non-bacterial and fungal reads were filtered from the respective datasets. To remove potential sequencing errors bacterial OTUs present in only 1 sample were removed and fungal OTUs with less than 5 counts were removed since unique OTUs had high relative abundances in their sample. Taxa that were highly abundant in negative controls in comparison to subject samples were identified as contaminants and removed with the Decontam package (v1.28.0).^45^ To achieve even sampling depths across samples, rarefaction was performed using the Vegan package (v2.7.2).^46^ The chosen sampling depths for bacterial and fungal datasets were 2900 and 1200 reads, respectively. Since the sampling depth of wound samples for bacterial reads was significantly greater than intact skin samples, a separate sample size of 23000 reads was chosen for bacterial analysis of wound samples alone.

Microbial diversity was evaluated by assessing relative abundance of taxa and beta and alpha diversity. To determine the relative abundance of taxa in each sample, phyloseq::tax_glom() was used to agglomerate OTUs to the genus level. Genera with less than 20 reads were removed and those not part of the top 15 most abundant were renamed to Other to reduce noise. Bray Curtis dissimilarity distances were calculated using phyloseq::distance() for beta diversity. Distances were ordinated using phyloseq::ordinate() for nonmetric multidimensional scaling (NMDS) and statistical differences between groups were tested with PERMANOVA using vegan::adonis2. Phyloseq::estimate_richness() calculated alpha diversity metrics, including Shannon index and observed OTUs, and Wilcoxon rank sum test determined statistical differences between groups. Visualizations were made with ggplot2 (v4.0.1) and ggpubr (v0.6.3).

The mean, standard deviation, range, and percentages of clinical variables shown in Table I were calculated within an R statistical environment. The Wilcoxon rank sum test was used to assess longitudinal changes in wound pH and volume. Univariable screening followed by multivariable regression was used to determine clinical variables correlated with wound pH. Variables that remained significantly correlated with pH remained in the final model. Pearson’s correlation was used to quantify the relationship between wound pH and volume.

**Table I.**
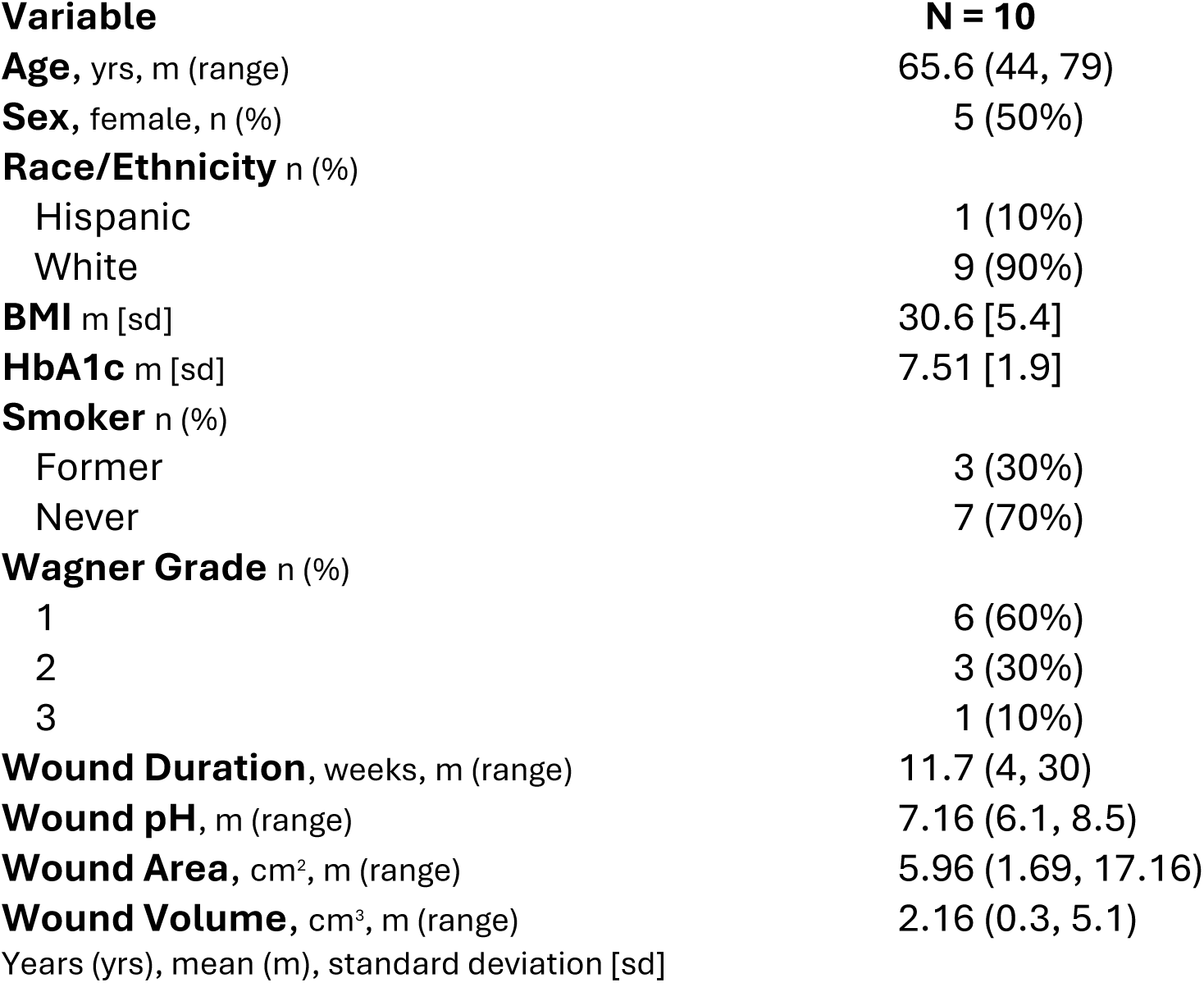
Cohort Characteristics.

| Variable | N = 10 |
| --- | --- |
| **Age**, yrs, m (range) | 65.6 (44, 79) |
| **Sex**, female, n (%) | 5 (50%) |
| **Race/Ethnicity** n (%) |  |
| Hispanic | 1 (10%) |
| White | 9 (90%) |
| **BMI** m [sd] | 30.6 [5.4] |
| **HbA1c** m [sd] | 7.51 [1.9] |
| **Smoker** n (%) |  |
| Former | 3 (30%) |
| Never | 7 (70%) |
| **Wagner Grade** n (%) |  |
| 1 | 6 (60%) |
| 2 | 3 (30%) |
| 3 | 1 (10%) |
| **Wound Duration**, weeks, m (range) | 11.7 (4, 30) |
| **Wound pH**, m (range) | 7.16 (6.1, 8.5) |
| **Wound Area**, cm<sup>2</sup>, m (range) | 5.96 (1.69, 17.16) |
| **Wound Volume**, cm<sup>3</sup>, m (range) | 2.16 (0.3, 5.1) |
| Years (yrs), mean (m), standard deviation [sd] |  |

## Results

### Study design and subject characteristics

10 subjects with a chronic DFU, defined as an ulcer present for >4 weeks that did not respond to standard of care, were enrolled and treated with a PGA/PLCL bioresorbable matrix. The baseline wound status was established at visit 1, prior to application of the matrix, and were re-evaluated over the following 4-week period, or until healed (**Figure 1**). To allow the hydroxy acids to fully absorb into the wound bed, wounds were reassessed and a new matrix was reapplied every 14 days at visits 3 and 5, along with standard of care. Changes to the DFU microenvironment were evaluated by assessing wound pH, the microbiome and inflammatory markers. At visits 2 and 4 subjects received standard of care without wound debridement. During the study, one subject was replaced due to an adverse event unrelated to the treatment that resulted in amputation of the study wound.

**Figure 1:**
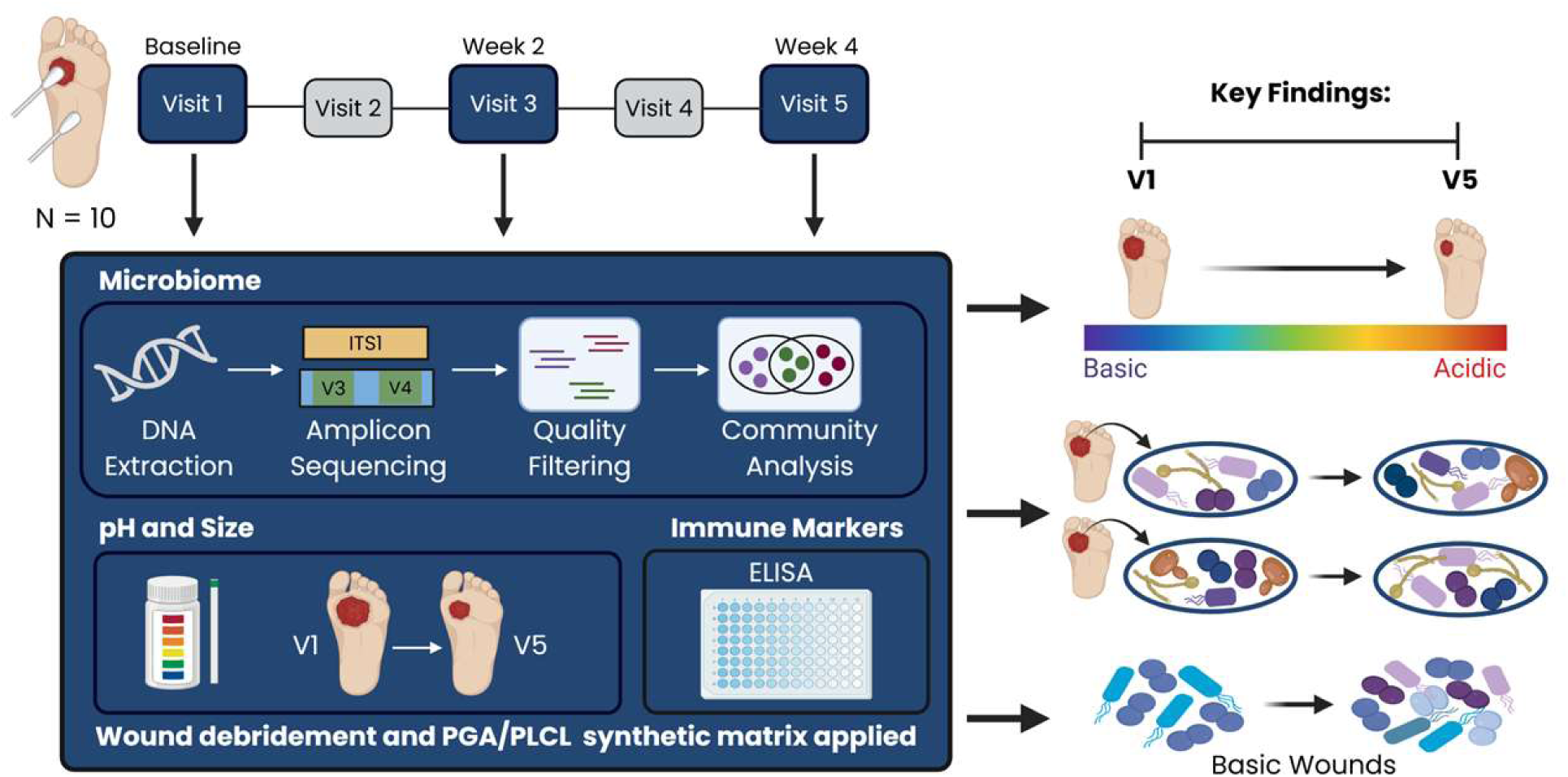
Overview of study to assess changes in the diabetic foot ulcer microenvironment during treatment with a PGA/PLCL synthetic matrix. 10 adults with a chronic DFU were treated with the PGA/PLCL wound matrix. Baseline data was collected at visit 1, prior to treatment, and follow-up visits to observe changes in DFU microenvironment occurred in 2-week intervals at visits 3 and 5 (n=5 visits). Wound pH and size were measured, and swabs were used to collect wound fluid for microbial and immune marker analysis. Swabs were also collected from intact skin to compare microbiomes. Wound debridement and application of the matrix occurred at visits 1, 3 and 5 as well. At visits 2 and 4 subjects received standard of care without debridement. During treatment the pH and volume of all wounds decreased. Individual shifts in the DFU microbiome were observed during treatment, as well as a trend where wounds with a basic pH at baseline increased in diversity. "Created in BioRender. Kalan, L. (2026) https://BioRender.com/a1042q8"

Subject demographics and baseline wound measurements are shown in **Table 1**. Wounds were located on the toes (n=3), plantar (n=3), lateral (n=1), medial (n=1), midfoot(n=1) and hindfoot(n=1) regions of the foot. The largest wounds were located on the midfoot, hindfoot and plantar regions with the smallest wounds located on the toes. During the study period most wounds were on a healing trajectory with 20% healed (healed n=2) and the remaining wounds ongoing (ongoing n=8). Wounds that healed had the lowest pH values (6.1 and 6.6) and smallest volumes (0.3cm^3^ and 0.8cm^3^) at baseline.

### Wound pH and volume

Wound pH and volume were measured at baseline and every 2-weeks throughout treatment. The average pH at visit 1, prior to treatment, was 7.16 (SD 0.7) and by visit 3 all wounds became acidic with an average pH of 5.69 (SD 0.56) (Wilcoxon rank sum test p<0.001). Wound pH further decreased to an average of 4.83 (SD 0.28) by visit 5 (Wilcoxon rank sum test p<0.001; **Figure 2A**). At baseline the average wound volume was 2.16 cm^3^ (SD 1.83) and decreased to 1.15 cm^3^ (SD 1.49) and 0.99 cm^3^ (SD 1.40) by visits 3 and 5, respectively. All wounds decreased in size during the treatment with a significant difference found by visit 5 (Wilcoxon rank sum test p<0.05; **Figure 2B**). Overall, all wounds decreased in pH and volume during treatment, demonstrating a healing trajectory.

**Figure 2:**
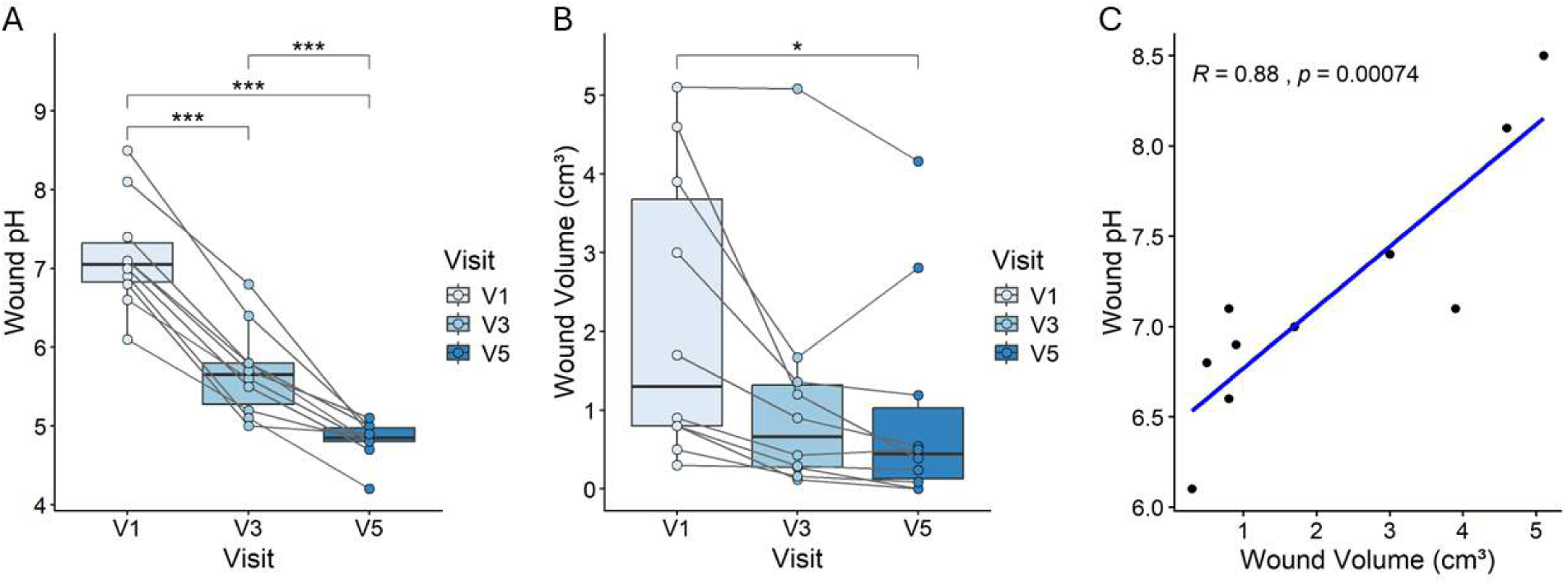
Wound pH and volume are positively correlated and decreased during treatment. A) Wound pH values measured prior to debridement at baseline (V1) and following treatment with the PGA/PLCL synthetic matrix (V3 and V5). Values from the same subject are connected by lines. Pairwise Wilcoxon rank sum tests were used to determine statistical significance between visits (V1 vs V3, V1 vs V5, and V3 vs V5). p<0.05 = *, p<0.01 = **, p<0.0001 = ***. B) Wound volume (cm^3^), measured post-debridement, at baseline (V1) and following treatment with the matrix. Wilcoxon rank sum tests were used to determine statistical significance. Only significant comparisons are shown. C) Pearson’s correlation (R = 0.88) was performed to test the relationship between wound pH and volume at the baseline visit. The blue line represents the linear regression.

Since wound pH has been correlated with wound outcomes in previous studies^16,17^, we aimed to determine if pH was related to other clinical variables. A linear model showed wound volume as the only variable correlated with wound pH (Pearson rho = 0.88; p<0.001; **Figure 2C**). The relationship revealed that larger wounds were more basic while smaller wounds were more acidic.

### Wound microbiome

We next assessed changes in the wound microbiome. Amplicon sequencing of the V3-V4 16S rRNA bacterial gene and ITS1 fungal region was performed on swabs collected from the wound bed and adjacent intact skin of 10 individuals at visits 1 (n=20), 3 (n=20) and 5 (n=16) as well as negative (n=19) and positive controls (n=1). After quality control and filtering, 12 samples across the V3-V4 16S and ITS1 datasets were removed from analysis due to poor sequencing depth. A total of 275 bacterial OTUs, which are representative of individual taxa, were detected across 28 wound and 26 intact skin samples and 393 fungal OTUs across 25 wound and 21 intact skin samples. At the genus level, the most represented bacteria identified in wounds were *Staphylococcus, Finegoldia*, *Corynebacterium* and *Anaerococcus* (**Figure 3A**). As for fungi, the most abundant genera were *Candida, Cladosporium, Alternaria* and *Aspergillus* (**Figure 3B**).

**Figure 3:**
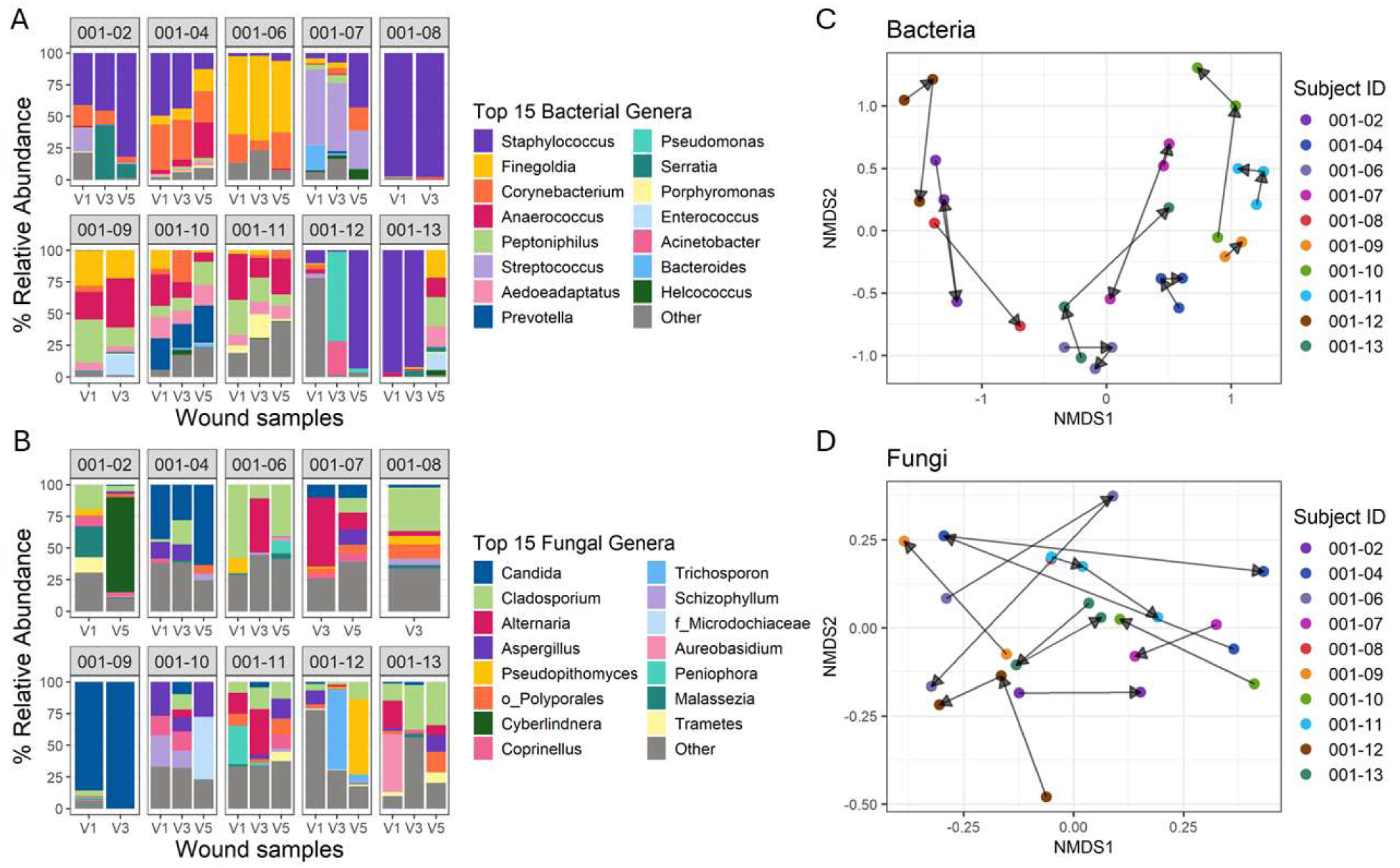
Wound-specific changes in bacterial and fungal communities during treatment. A) Relative abundance of the top 15 bacterial genera in wounds for each subject at each visit. Taxa outside of the top 15 were agglomerated into Other. B) Relative abundance of top 15 fungal genera in each wound sample. Taxa that were not classified to the genus level are represented by the family (f_) or order (o_) name. C) Non-metric dimensional scaling (NMDS) ordination of Bray-Curtis distances calculated between bacterial communities from wound samples. Arrows represent the shift in bacterial communities from V1 to V3 to V5 for each subject. Samples positioned closer together are more similar in community composition than samples positioned farther apart. PERMANOVA determined statistical significance of community composition between subjects (p<0.001, R^2^ = 0.6817). D) Shifts in wound fungal communities for each subject visualized with NMDS ordination of Bray-Curtis distances. The arrows represent the shift in fungal composition between visits for each subject. PERMANOVA determined statistical significance of community composition between subjects (p<0.001, R^2^=0.50742).

To understand how the PGA/PLCL matrix may have impacted the microbiome, we assessed changes in the bacterial and fungal communities over the study. Alpha diversity metrics were used to measure microbial diversity within each sample, including the observed number of OTUs and Shannon’s index, a measure of the number of OTUs and how evenly distributed they are in a sample. No significant differences in bacterial or fungal diversity were observed across visits (**Supp. Figure 1**). Beta diversity was measured by calculating Bray-Curtis dissimilarity to evaluate differences in overall community composition between samples. Bray-Curtis metrics use the presence or absence and relative abundance of each taxon to determine how similar two samples microbiomes are to each other. Samples positioned closer together in NMDS ordination have more similar microbiomes whereas those positioned farther apart are more dissimilar, indicating greater differences in microbial composition. A significant difference in bacterial and fungal composition was found between samples from different subjects, indicating the wound microbiome had strong subject specificity (**Figure 3C** bacteria: PERMANOVA p<0.001, R^2^ = 0.6817; **Figure 3D** fungi: p <0.01, R^2^=0.50742). Shifts in bacterial and fungal communities for each subject from visit to visit are represented by the length of arrows in the NMDS plots. Changes in the microbial community structure for individual subjects across visits (i.e., over time) was observed, however a consistent shift in the same direction was not apparent, suggesting microbial community structures change over time in a wound-specific manner.

Since wound pH progressively decreased towards an acidic, intact skin-like pH, we investigated if the microbiome became progressively similar to intact skin. Beta diversity was used to determine if microbial communities of the wound and intact skin were distinct. No statistical difference was identified when considering bacterial composition (PERMANOVA p>0.05, R^2^=0.0322; **Suppl. Figure 2A**) indicating bacterial communities were similar between sample types. Fungal composition of wound and intact skin, however, were significantly different (PERMANOVA p<0.05, R^2^=0.0386; **Suppl. Figure 2B**), although modest separation in NMDS ordination was observed indicating the communities share taxa. This suggests that despite wound acidification and potential differences in pH, the microbiomes of intact skin and wound were not clearly distinct.

We next evaluated associations between baseline wound pH and bacterial community structure. When assessing beta diversity, the dissimilarity between microbial communities of different samples, wounds that were basic or acidic at baseline clustered by pH grouping indicating that wounds with the same starting pH had similar bacterial composition. Although, this trend was insignificant (PERMANOVA p>0.05, R^2^ = 0.10244; **Figure 4A**). We then evaluated differences in alpha diversity (within sample diversity), measured using the Shannon index, for each subject across each visit. This revealed that basic wounds at baseline (n=5) had an overall increase in bacterial diversity over time, while wounds with an acidic starting pH (n=4) had less change in diversity (**Figure 4B**). Furthermore, the basic baseline pH group had an overall greater Shannon index in comparison to wounds with an acidic baseline pH (Wilcoxon rank sum test p<0.05; **Figure 4C**). Only 1 subject had a neutral wound pH at baseline preventing statistical analysis. The overall changes in alpha diversity suggest that bacterial communities may be impacted differently by micro-acidification from the matrix dependent on the baseline pH.

**Figure 4:**
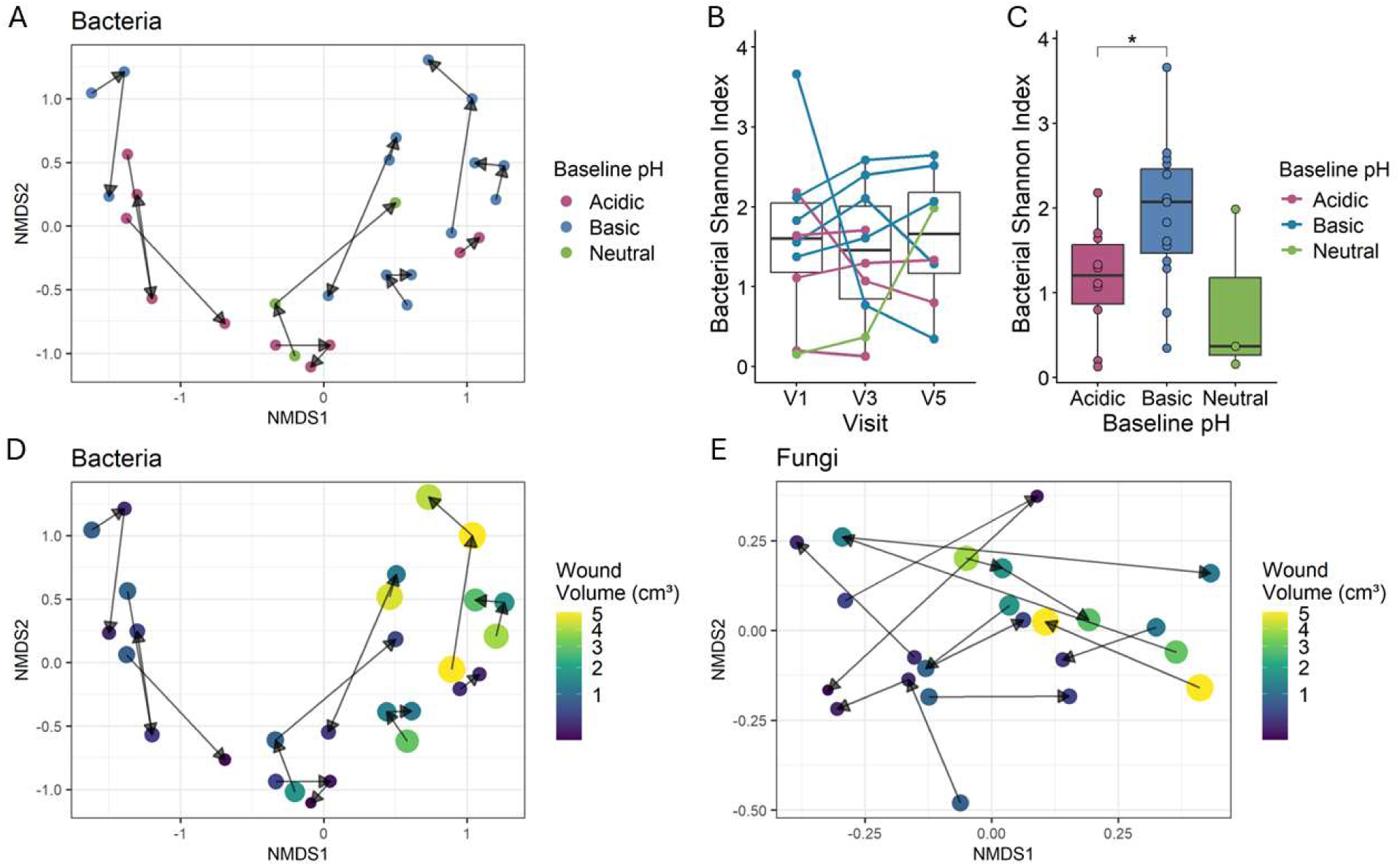
Microbial communities were associated with baseline pH and size. A) NMDS ordination of Bray-Curtis distances assessing differences in wound bacterial composition between baseline pH groups. PERMANOVA found insignificant separation between wounds with a basic (n=5), acidic (n=4) or neutral (n=1) pH at baseline (p>0.05, R^2^ = 0.10244), however wounds with same pH at baseline trend to cluster together. B) Changes in bacterial diversity, measured by Shannon’s index, between visits for each subject are represented by each line. Subjects are coloured by their pH at baseline. C) Comparison of bacterial diversity, measured by Shannon index, between wounds with an acidic, basic or neutral pH at baseline. Statistical differences were calculated with the Wilcoxon rank sum test. D) NMDS plot of Bray-Curtis distances calculated between bacterial communities from wound samples. PERMANOVA determined statistical significance in community composition based on wound volume (p<0.01, R^2^= 0.11557). Larger and lighter coloured points represent wounds with larger volumes. E) Differences in fungal communities in wound samples visualized with NMDS ordination of Bray-Curtis distances. PERMANOVA determined statistical significance of community composition based on wound volume (p<0.05, R^2^= 0.06942).

Given the association between wound pH and volume, we next investigated if there is an association between wound volume and microbiome composition. We found that larger wounds had more similar bacterial composition to each other than to smaller wounds, since wounds of a larger size clustered together when assessing beta diversity (PERMANOVA p<0.01, R^2^= 0.11557; **Figure 4D**). Fungal communities showed the same trend and resulted in significant separation by wound volume (PERMANOVA p<0.05, R^2^= 0.06942; **Figure 4E**).

### Immune profile in wounds

Since DFUs can stall in an inflammatory state, we wanted to determine if altering wound pH with the PGA/PLCL synthetic matrix would affect the surrounding immune profile. ELISA was used to quantify the concentration of a panel of cytokines, chemokines, and growth factors. Concentrations of analytes were highly variable between subjects, and none of the measured immune markers showed a consistent trend across all subjects during the treatment period (**Supplemental Table 1**). Focusing on larger wounds, we identified elevated levels of the pro-inflammatory chemokines monocyte chemoattractant protein-1 (MCP-1) and macrophage inflammatory protein-1 δ (MIP-1δ) (**Figure 5**). With MCP-1 we specifically observed a trend where levels increased between visits 1 and 3 (**Figure 5A**).

**Figure 5:**
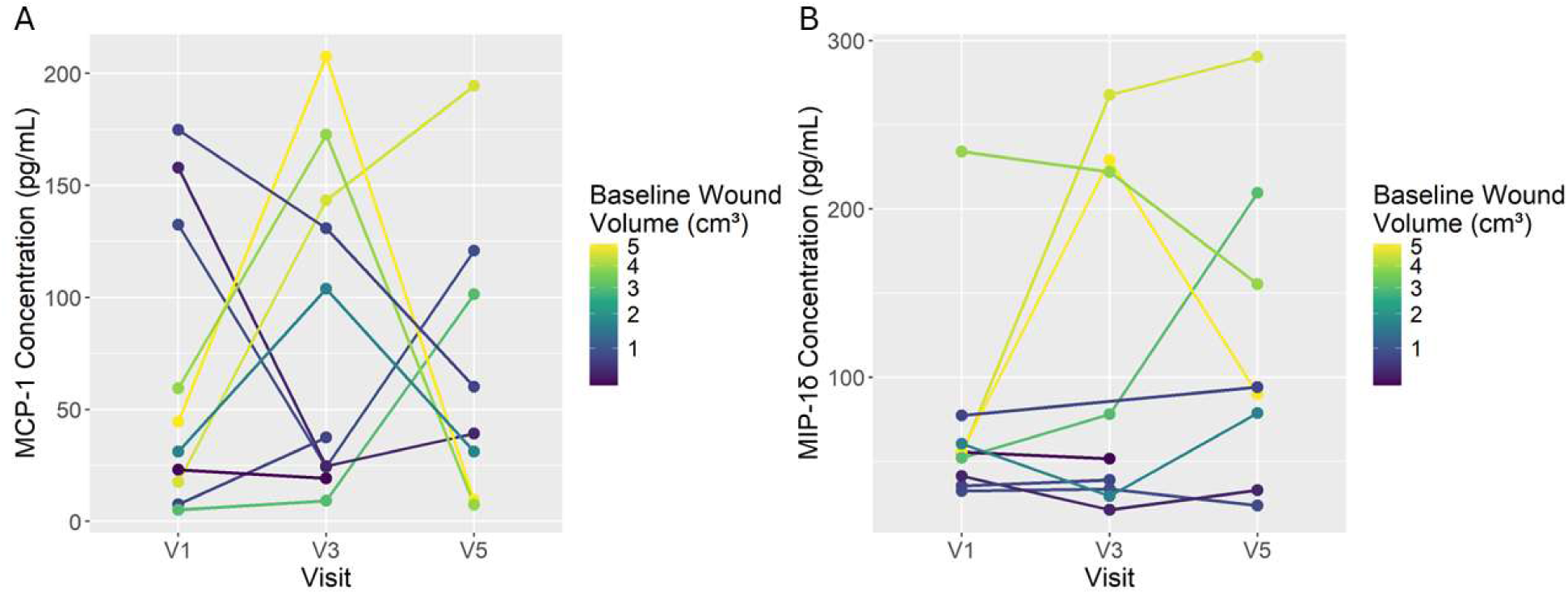
Larger wounds were elevated in the pro-inflammatory chemokines MCP-1 and MIP-1δ. A) Enzyme linked immunosorbent assay was used to determine the concentration of MCP-1 in wounds and values were normalized to total protein concentration. Lines coloured by wound volume at baseline represent the changes in MCP-1 concentration between visits for each subject. B) Lines coloured by wound volume at baseline represent the changes in MIP-1δ concentration between visits for each subject.

Acid-releasing dressings have emerged as a therapeutic strategy that can act to perturb the stalled alkaline and pro-inflammatory microenvironment in DFUs and promote healing.^6,47^ While wound pH can affect the growth of colonizing microbiota, the effects of reducing wound pH on the microbiome remain poorly understood. Here, we treated 10 DFUs with a PGA/PLCL synthetic matrix to evaluate the effects of wound pH on the microbiome and immune profile.

Compared to baseline, we found that all wounds in the study appeared to be on a healing trajectory and showed a decrease in both pH and wound volume. Moreover, 2 wounds achieved complete closure in the 5-week study period. We found that pH and wound volume are positively correlated with larger wounds having a more basic pH at baseline. As pH decreased throughout treatment, we identified wound-specific shifts in both bacterial and fungal communities, which may suggest that altering pH can result in changes to microbial communities respective to their baseline structure. Our findings also revealed a trend of increasing bacterial diversity as pH became progressively acidic in wounds that had a basic pH prior to treatment. In contrast, wounds that had an acidic pH at baseline remained relatively stable in bacterial diversity over time, which may be related to the maintenance of an acidic microenvironment in comparison to the transition from basic to acidic. Furthermore, wounds of a similar size and those from the same pH group at baseline were more similar in bacterial and fungal composition, suggesting wound size and microenvironment may contribute to microbial community structure. Larger wounds were also elevated in the pro-inflammatory chemokines MCP-1 and MIP-1δ, both of which contribute to inflammation through monocyte recruitment but also have important roles in angiogenesis and fibroblast migration, respectively.^48,49^ In summary, we found that a higher baseline pH was associated with increasing bacterial diversity and inflammation was driven by wound size.

This work revealed a relationship between wound size and the microenvironment where larger wounds were more basic, inflammatory and had similar microbial composition. Previous clinical studies have similarly found a positive correlation between pH and area in DFUs as well as other wound etiologies.^16,50,51^ Recent work has also found that DFUs with a higher pH are correlated with longer wound durations and increased necrotic tissue, further demonstrating that more alkaline environments are associated with wound severity.^16,28^ Previous studies have also explored the relationship between the prevalence of bacterial taxa and wound pH using culture based and targeted PCR methods. Shukla et al.^29^ cultured isolates from 50 wounds of mixed etiology and found an association of *Pseudomonas aeruginosa* with pH >8.5, *E. coli*, *Klebsiella pneumoniae* and *Proteus* with pH 8 and *S. aureus* with pH 7.5, which is consistent with the established view that alkaline environments support the growth of pathogens. Similar to our findings, other studies, including one which sampled 100 DFUs, did not find a relationship between specific bacterial taxa and wound pH.^28,30^ Here, the observed increase in bacterial diversity in wounds that transitioned to an acidic pH may suggest a greater community level shift due to the changing microenvironment. However, future work with a larger sample size or study period may be able to detect specific changes in the relative abundance of taxa.

Apart from community level shifts, metabolic changes of wound microbiota could have occurred as well in response to acidification. Future work could explore transcriptional changes to better understand how microbial growth, metabolism, and virulence differ in basic and acidic wound conditions and its connection to wound outcome. For example, in basic conditions, some *Candida* species preferentially grow as hyphae and produce thicker biofilm, but in acidic conditions, this phenotype is suppressed and growth as yeast is upregulated.^26,27^ While wound-specific changes occurred in fungal communities, the complex pH regulatory systems which are common in fungi could explain why more trends were observed between bacteria and pH. Thus, assessing functional changes in response to acidification could give more insight into the effect of pH on the wound microbiome and healing.

Previous studies have shown that wound acidification can polarize M1 pro-inflammatory macrophages towards an M2 anti-inflammatory phenotype to restore the M1/M2 imbalance and improve healing.^22,52^ Based on our analysis of the DFU immune profile and limited array of immune markers, we were not able to characterize the wound environment as pro- or anti-inflammatory and were only able to detect trends associated with levels of chemokines. MCP-1 and MIP-1δ may have been elevated in larger wounds since they likely have more inflammatory stimuli. While both chemokines are considered pro-inflammatory and are critical for the inflammatory phase of healing, they also have a role in the proliferative phase by promoting angiogenesis and fibroblast migration.^48,49^ This activity may be related to the large amount of wound closure as larger wounds decreased substantially in volume over time.

The results of this work were limited by the small sample size. Subject specificity has a strong effect on the microbiome due to variation between individuals, which likely limited the ability to detect specific longitudinal changes. A larger cohort could have been beneficial to better isolate potential differences in basic, acidic and neutral wounds. Furthermore, with a larger cohort, the effect of acidification on different wound severities could help better resolve the effects of pH on healing trajectory. The analysis was also limited to genus level differences since sequencing the V3- V4 16S rRNA and ITS1 regions is not able to provide confident species-level resolution. Furthermore, to better assess the efficacy of pH-reducing materials and the effect of wound acidification, future work could compare this treatment to current standard of care or other wound matrices.

Overall, this study was able to define trends between wound pH, microbiome, and inflammation in DFUs. As critical components to the DFU microenvironment, it is important to understand the influence of pH on wound microbiota and inflammation in healing to grasp the full therapeutic potential of pH-reducing materials. This work supports the possibility of wound acidification as an integrated treatment approach that targets multiple components of the complex DFU microenvironment to improve wound outcomes.

## Key Findings

- Wound pH continuously and significantly decreased during application of a PGA/PLCL synthetic matrix to the wound bed
- 8 out of 10 wounds were on a healing trajectory after application of the PGA/PLCL synthetic matrix.
- Wound-specific shifts in microbial communities occurred during treatment with more alkaline wounds increasing in bacterial diversity.
- Wound volume was significantly associated with bacterial and fungal composition and larger wounds were elevated in pro-inflammatory chemokines, MCP-1 and MIP-1δ.

## Acknowledgements

We would like to acknowledge the McMaster University Genomic Center and Surrette lab for amplicon sequencing services. We also acknowledge members of the Kalan laboratory for feedback and discussion.

## Statements and Declarations

### Ethical Considerations

This study was approved by the WCG Institutional Review Board for activities at the PA Foot and Ankle Associates and the Hamilton Integrated Research Ethics Board for activities at McMaster University

### Consent to Participate

All subjects participating in the study provided informed consent in writing.

### Consent for Publication

Not applicable

### Data Availability

The NCBI BioProject number for this project is PRJNA1514187. Code for analysis and figure generation can be found on GitHub at: https://github.com/Kalan-Lab/Effect_of_reducing_pH_on_DFU_microenvironment

## Abbreviations and Acronyms

DFU: Diabetic foot ulcer
ELISA: Enzyme linked immunosorbent assay
ITS1: Internal transcribed spacer 1
MCP-1: Monocyte chemoattractant protein 1
MIP-1δ: Monocyte inflammatory protein 1 δ
NMDS: Nonmetric multidimensional scaling
OTU: Operational taxonomic unit
PGA/PLCL: Polyglycolic acid/poly(lactide-co-caprolactone)
QIIME2: Quantitative insights into microbial ecology 2

