## Supplemental Figures for "The Effects of Reducing pH with a Bioresorbable Synthetic Matrix on the Diabetic Foot Ulcer Microenvironment"

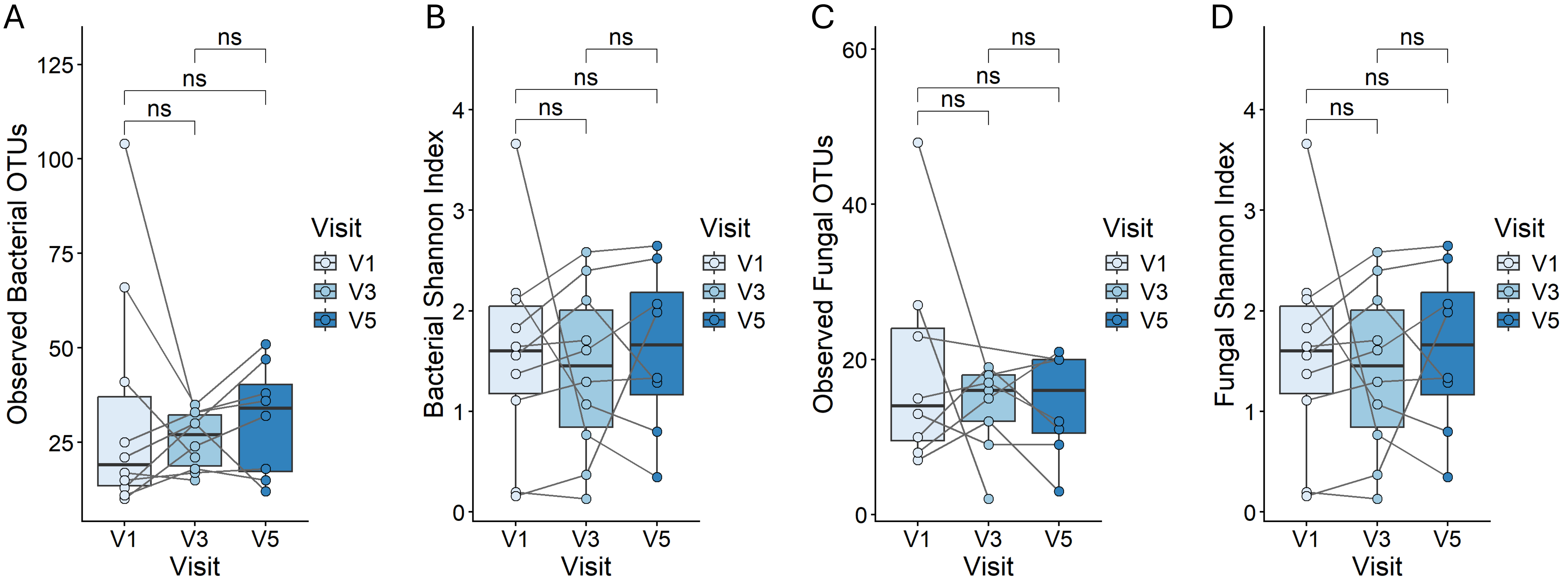


**Supplemental figure 1: No changes in alpha diversity were found in bacteria or fungi during treatment.** A) Alpha diversity, measured by the observed number of OTUs, was used to assess bacterial diversity at each visit. Subjects are connected by lines. Wilcoxon rank sum test did not find statistical significance. B) Shannon index was used to measure alpha diversity of bacterial communities during treatment. No significant differences were identified by Wilcoxon rank sum test. C) Fungal alpha diversity was measured by the observed number of OTUs. Wilcoxon rank sum test did not find statistical significance. D) Insignificant differences were found in Shannon's index of fungal communities between visits by Wilcoxon rank sum test.

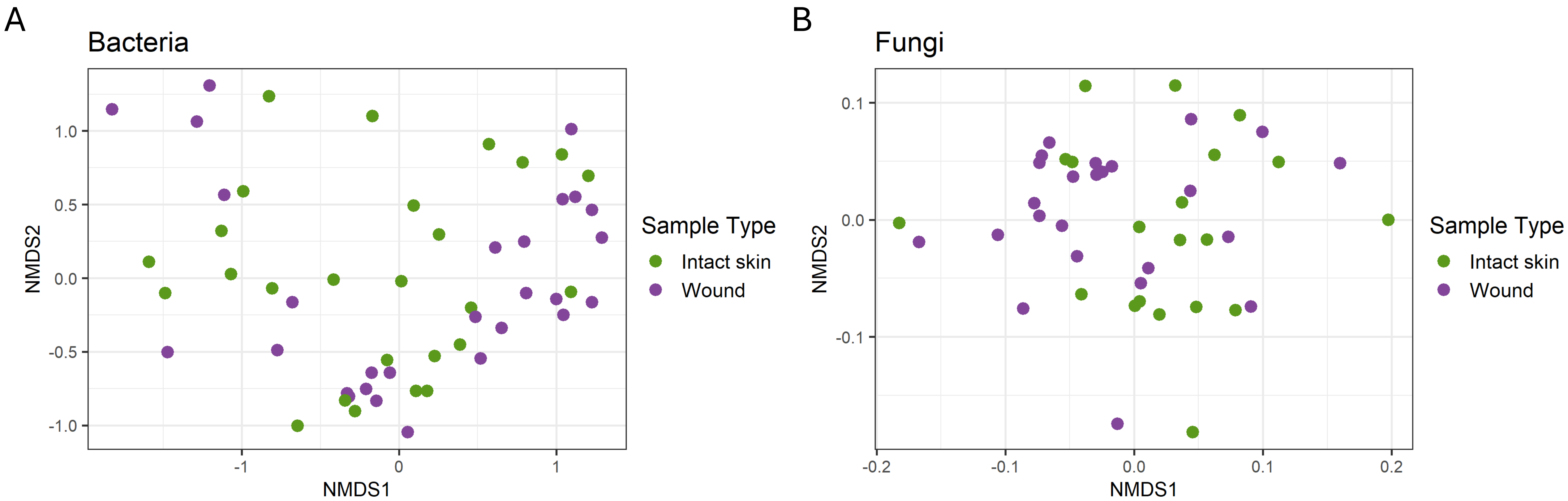


**Supplemental figure 2: Microbial communities in wounds and intact skin are similar** A) NMDS plot of Bray-Curtis distances calculated between bacterial communities from wound and intact skin samples. PERMANOVA found insignificant separation between wound and intact skin ($p>0.05$, $R^2=0.0322$. B) Differences in fungal communities in wound and intact skin samples visualized with NMDS ordination of Bray-Curtis distances. PERMANOVA determined statistical significance of community composition between wound and intact skin ($p<0.05$, $R^2= 0.0386$), however visual separation is modest.
